# Monkeypox in Europe: Epidemiology and Risk Factors – A Scoping Review Study

**DOI:** 10.1101/2024.08.28.24312706

**Authors:** Nandakumar Ravichandran, Parnian Jalili

**Affiliations:** School of Medicine, University College Dublin, Belfield, Dublin 4, Ireland; School of Public Health, University College Dublin, Belfield, Dublin 4, Ireland

**Keywords:** Monkeypox, epidemiology, infectious, neglected tropical disease, public health

## Abstract

**Background:** Monkeypox (mpox) is a zoonotic disease originating from the Congo Basin (Clade I) and West Africa (Clade II). In 2022, mpox spread to non-endemic European countries, predominantly through human transmission associated with sexual contact. The outbreak in Europe was primarily with the Clade IIb lineage, which is less virulent. The World Health Organization (WHO) declared this outbreak a Public Health Emergency of International Concern (PHEIC) in 2022, which ended in May 2023 after a decline in cases. However, in July 2024, a resurgence of the more virulent Clade I occurred in the Democratic Republic of Congo (DRC), leading WHO to declare mpox a PHEIC again, due to the risk of global spread. Understanding epidemiology and risk factors of mpox is vital for effective public health measures.

**Methodology and principal findings:** A search conducted from 2014 to 2024 across PubMed, Scopus and Embase identified 38 studies on mpox in Europe, which were included for qualitative analysis. The key themes identified were epidemiology and risk factors/ behaviors. High-risk behaviors include sexual contact among men who have sex with men (MSM) with multiple partners, living with HIV, and frequent travel to endemic regions.

**Conclusions and significance:** With no definitive cure for mpox, public health measures such as surveillance, monitoring, and contact tracing are essential. Additionally, encouraging case-control studies is crucial for exploring other potential risk behaviors and design behavioral interventions, vaccination campaigns and awareness programs aimed at reducing high-risk behaviors among these populations. Although the number of cases in Europe did not surge in August 2024, proactive measures are necessary to prevent further spread.

## Introduction

Monkeypox (mpox) is a neglected tropical disease caused by the Monkeypox virus (MPXV), a double-stranded DNA virus belonging to the Orthopoxvirus (OPXV) genus of the Poxviridae family. The virus was first identified in 1958 during an outbreak among Cynomolgus monkeys at the Statens Serum Institute in Copenhagen, Denmark (1). Although initially discovered in a laboratory setting, the virus is believed to originate from the tropical rainforests of the Congo Basin and West Africa. Based on genetic and geographical analyses, MPXV is classified into two main clades: Clade I, associated with the Congo Basin, and Clade II, which originates from West Africa. These clades differ by approximately 900 base pairs in genomic length (2). Clade I is more virulent, with a mortality rate of up to 10%, while Clade II, which includes subclades IIa and IIb, generally has a lower mortality rate, with Clade IIa exhibiting less than 1% mortality. Clade IIb, in particular, is notable for its higher human-to-human transmission rates. A 2023 animal model study demonstrated that mice survived even when exposed to the highest dose of the Clade IIb.1 strain. The study observed virus replication in the lungs and abdominal organs, with virulence and lethality ranked as follows: Clade I > Clade IIa > Clade IIb.1, suggesting that Clade IIb.1 is less virulent and deadly (3).

Mpox manifests with a range of symptoms, including skin rashes (vesicular), fever, headaches, fatigue, swollen lymph nodes (lymphadenopathy), mucosal lesions, and genital rashes. The characteristic rash typically begins as a flat sore that develops into a blister, which then turns into a lesion, dries up, crusts over, and eventually falls off. The rash usually starts on the face and spreads across the body, including the palms and soles of the feet (4). The transmission of mpox primarily occurs through close contact with an infected person’s mucosal lesions, droplets, and face-to-face interactions. Additionally, contaminated materials (such as bedding or clothing) and sexual contact can also facilitate the spread of the virus. However, there is no evidence to support airborne transmission of mpox (5). Although the disease was first recognized in humans in 1970, when it was identified in a nine-month-old infant in Zaire (now the Democratic Republic of Congo (DRC)) (6), the virus’s pathogenic potential was already highlighted in 1987 as a significant OPXV affecting humans (7).

The first outbreak of mpox outside Africa occurred in the United States in 2003, linked to the importation of animals from Ghana, particularly African rodents (2, 8). These rodents infected pet prairie dogs, which subsequently transmitted the virus to humans, including a three-year-old girl in Central Wisconsin. This outbreak was caused by Clade II, a less virulent strain than those found in the Congo Basin (9).

In 2005, South Sudan reported another outbreak, where the isolated virus shared the phylogenetic characteristics of the Congo Basin’s Clade I, supporting the theory of virus importation from the Congo Basin and underscoring the potential for person-to-person transmission (10). Subsequently, in 2017-18, Nigeria experienced a mpox epidemic, during which sexual transmission was documented in several patients (11). In 2020, the United Kingdom reported a travel-associated case on May 7th in a patient who had returned from Nigeria (12).

From January 2022 to June of the same year, an increasing number of mpox cases were reported in non-endemic countries across Europe. The overall Case Fatality Rate (CFR) in Europe during this period was reported at 3.6%, with higher mortality observed among children, young adults, immunocompromised individuals, and cases linked to sexual contact (8).

Although mpox was considered a rare viral disease by the World Health Organization (WHO) until 2018, in July 2022, the WHO declared it a Public Health Emergency of International Concern (PHEIC) due to its rapid spread via sexual contact to non-endemic countries, posing a high risk in Europe and a moderate risk worldwide. In May 2023, the PHEIC was declared over after a sustained decline in cases globally. However, there was a resurgence in cases in the DRC last year with the spread of a new virus strain, Clade Ib. Since July 2022, cases of clade Ib have been reported in neighboring countries of DRC, and the WHO declared the disease a PHEIC once again on August 14^th^, as the current strain is more virulent stating its potential to spread across countries in Africa and outside the continent (13, 14). On August 15^th^, Sweden reported a case of Clade Ib who returned from Africa (15).

Given these developments, there is an urgent need to study the epidemiology and risk factors associated with mpox. Such research will be crucial for designing future studies and planning interventions to curb the spread of the disease. This scoping review aims to analyze and report on the epidemiological trends and risk factors related to mpox spread in Europe.

## Methods

This scoping review followed Arksey and O’Malley’s six-step methodological framework (16), incorporating additional recommendations by Levac et al. (2010) (17). The process included identifying the research questions, conducting comprehensive search, selecting relevant studies, charting data, and summarizing the findings.

### Step 1 – Identifying the research question

The research question identified was “What are the risk factors and associated behaviors contributing to Mpox transmission and prevalence in Europe, based on current epidemiological evidence?”

### Stage 2 – Identifying relevant studies

A search was conducted over the last 10 years (2014-2024), focusing on English-language publications within the PubMed, Scopus and Embase databases. The search syntax employed in this review is

**((“prevalence”[tiab] OR “epidemiology”[tiab] OR “Epidemiology”[MeSH]) AND (“Monkeypox”[tiab] OR “Monkey pox”[tiab] OR “Mpox”[tiab] OR “Monkeypox virus”[MeSH]) AND (“Europe”[tiab] OR “European region”[tiab] OR “Europe”[MeSH] OR “Western Europe”[tiab] OR “Eastern Europe”[tiab] OR “Northern Europe”[tiab] OR “Southern Europe”[tiab] OR “European Union”[tiab] OR “EU”[tiab] OR “EU countries”[tiab] OR “EU member states”[tiab]))**

### Stage 3 – Study selection

The initial search yielded 570 papers. The study selection process is outlined in the PRISMA flow chart (see Figure 1). Studies were included irrespective of their design. Web-based collaboration software was used for screening, citation management, and duplication removal.

**Figure 1.**
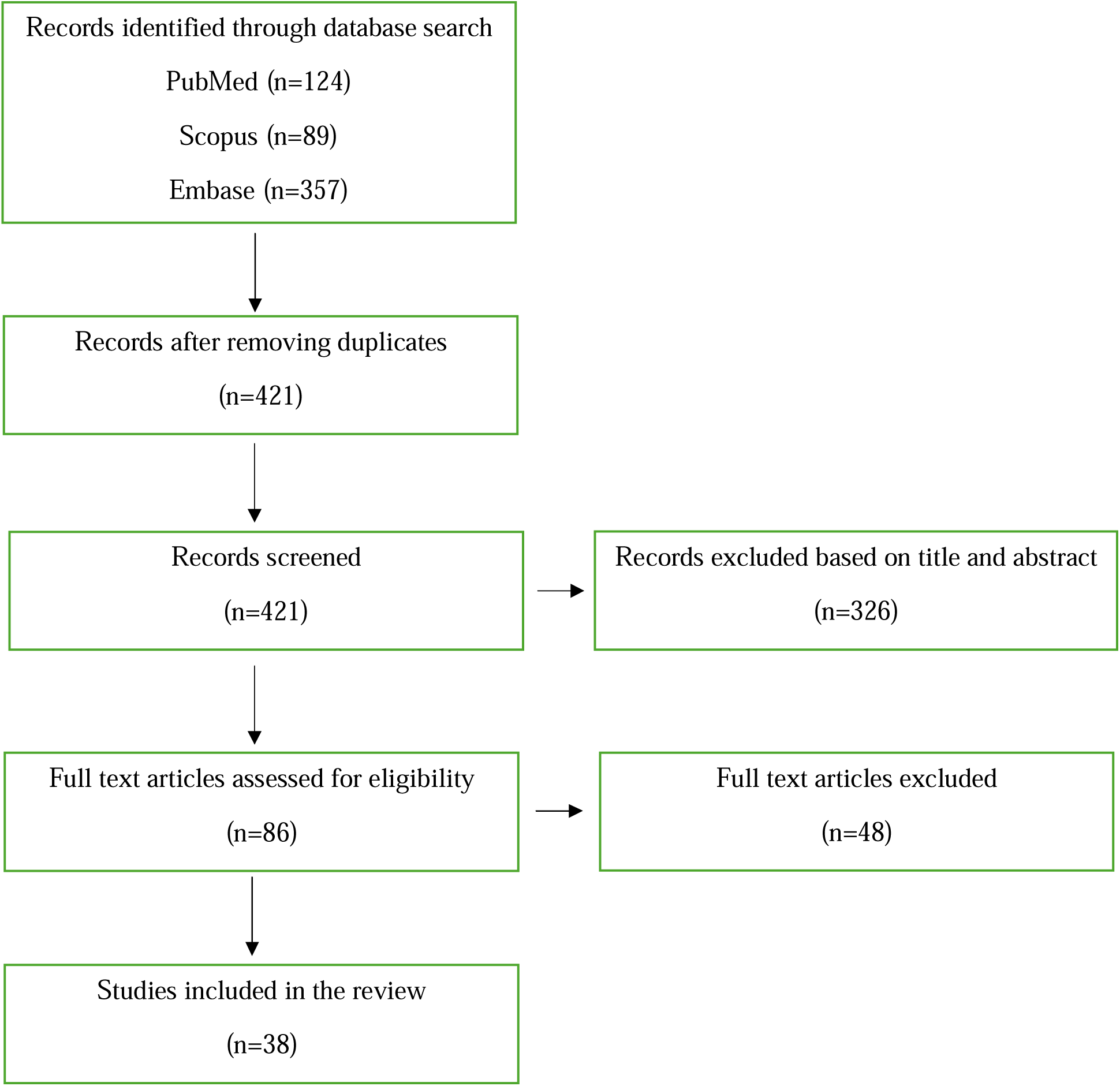
PRISMA Flowchart of the review.

#### Inclusion Criteria

- Focused on the prevalence and epidemiology of mpox within the European region.
- Studies conducted from 2014 to 2024
- Studies published in English or other languages, provided that a translated version is available online.

#### Exclusion Criteria

- Studies that are not focused on the European region.
- Studies address the pathophysiology or therapeutic interventions of mpox, rather than its prevalence and epidemiology.
- Narrative reviews, Systematic reviews and meta-analyses

After screening, 38 papers were selected for inclusion.

### Stage 4 – Charting the data

Data from the 38 selected studies were charted and summarized in a table format for qualitative synthesis. The table included the following information: Author, Year of Publication, Study Design, Study Population, and Major Findings.

### Stage 5 – Collecting and summarizing data

Extracted data were presented in Table 1. Key themes were identified through thematic analysis based on Braun and Clarke’s framework (18).

**Table 1.** Description of the studies included in the scoping review.

| Author(s), Year | Journal | Location | Aim/Topic | Study design | Major Findings |
| --- | --- | --- | --- | --- | --- |
| **Meo SA and Klonoff DC (2022)** | European Review for Medical and Pharmacological Sciences | Global | Human monkeypox outbreak: global prevalence and biological, epidemiological and clinical characteristics – observational analysis between 1970-2022 | Retrospective study | Between May 7, 2022, and July 1, 2022, majority of these cases are found in non-endemic countries (n=52) predominant in UK, Germany, Spain, France, Portugal and the Netherlands. |
| **Vallejo-Plaza A et al. (2022)** | Euro Surveillance | Spain | Mpox (formerly monkeypox) in women: epidemiological features and clinical characteristics of mpox cases in Spain, April to November 2022. | Retrospective study | Spain accounted for one third of women Mpox cases in Europe. Around 10% of the women cases in Spain were linked to a Tattoo studio. Higher complication rates were reported in women than in men. Significant differences between men and women cases who were HIV positive (40.8% vs. 4.4%) |
| **De Vos E et al. (2024)** | International Journal of Infectious Diseases | Belgium | Potential determinants of the decline in mpox cases in Belgium: A behavioral, epidemiological and seroprevalence study. | Cohort study | Major route of transmission – sexual contact, decline in cases in Belgium due to behaviour modification – reduced sexual partners and reduced attending events and self-diagnosis with the symptoms during the end of the epidemic. |
| **Vaughan AM et al. (2024)** | Euro Surveillance | European Region | Continued circulation of mpox: an epidemiological and phylogenetic assessment, European Region, 2023 to 2024 | Retrospective study | Period 1 covered week 10 of 2022 to week 22 of 2023; Period 2 covered week 23 of 2023 to week 21 of 2024. After the peak in July 2022, cases rapidly declined and remained low until a mild resurgence from June 2023. Cases reported in Period 2 were less than Period 1 and resurgence observed mainly in Spain, Germany, Portugal, UK and France. Most of the cases were male and MSM. 8 |
|  |  |  |  |  | of the 10 cases died reported to be living with HIV. Majority of them were infected with Clade IIb which is less virulent. |
| **Vusirikala A et al. (2022)** | Emerging Infectious Diseases | England | Epidemiology of Early Monkeypox Virus Transmission in Sexual Networks of Gay and Bisexual Men, England, 2022. | Retrospective study | 82 out of 85 cases are suspected to be in GBMSM sexual networks. 26% of cases living with HIV and 91% receiving PrEP. Cases reported attending sex-on-premises venues, festivals, private sex parties, or cruising grounds 21 days before symptom onset. |
| **Kinoshita R et al. (2023)** | Epidemiology & Infection | Europe and America | Impact of airline network on the global importation risk of mpox, 2022. | Modelling study | Large PV with closely connected flights facilitates transmission of infection (e.g. London, UK) |
| **Noe S et al. (2023)** | Infection | Germany | Clinical and virological features of first human monkeypox cases in Germany. | Retrospective study | First two cases identified in Germany reported to have homosexual intercourse, one of them was an MSM sex worker. |
| **Vaughan AM et al. (2022)** | Euro Surveillance | WHO European Region (41 countries) | A large multi-country outbreak of monkeypox across 41 countries in the WHO European Region, 7 March to 23 August 2022. | Retrospective study | Most of the cases were MSM and 93.9% reported transmission through sexual contact. 37.2% were HIV-positive. Other routes of transmission include non-sexual, non-mother-to-child and non-healthcare associated, or fomites (0.2%; 11/6,797). Only 16.8% of cases reported on smallpox vaccination. |
| **Valentino M et al. (2023)** | International Journal of Dermatology | Malta | Clinical features and diagnostic challenges of mpox (monkeypox) outbreak in Malta: a retrospective cohort study | Retrospective cohort study | Hostility of Malta favored risk behaviors - sexual contact with more than one partner three months prior to the Mpox diagnosis, group sex, chemsex and unprotected sex. 27% of cases were living with HIV. |
| **de Vries HJ et al. (2023)** | Euro Surveillance | Sweden | Mpox outbreak among men who have sex with men in Amsterdam and Rotterdam, the Netherlands: no evidence for undetected transmission prior to May 2022, a retrospective study. | Retrospective study | No indication of extensive undetected transmission of mpox among MSM in Netherlands before May 2022. |
| **Schuele L et al. (2024)** | Journal of Medical Virology | Netherlands | Circulation, viral diversity and genomic rearrangement in mpox virus in the Netherlands during the 2022 outbreak and beyond. | Retrospective study | Phylogenetic analysis suggests that several of the B.1 lineage clusters were introduced into the Netherlands and then spread locally in the community. |
| **McFarland SE et al. (2023)** | Euro Surveillance | Germany | Estimated incubation period distributions of mpox using cases from two international European festivals and outbreaks in a club in Berlin, May to June 2022. | Retrospective study | Cases identified reported attending Club C (Berlin, Germany), the gay pride festival (Gran Canaria, Spain) and the fetish festival (Antwerp, Belgium) in considerable rates. |
| **Candela C et al. (2023)** | Viruses | Italy | Human Monkeypox Experience in a Tertiary Level Hospital in Milan, Italy, between May and October 2022: Epidemiological Features and Clinical Characteristics. | Retrospective study | Cases reported as use of recreational substances – smoking, alcohol, chemsex drugs. Only 14% were vaccinated against smallpox in childhood. High risk behaviors reported as living with HIV (47%), history of STIs, MSPs, attended International Pride parades and large gatherings and travel history. |
| **Hoffmann C et al. (2023)** | HIV Medicine | Germany | Clinical characteristics of monkeypox virus infections among men with and without HIV: A large outbreak cohort in Germany. | Cohort study | All cases were MSM. 39 cases acquired infection from Grand Canaria event in Spain. 46.9% of cases living with HIV and only 47.6% of cases didn't report history of STIs in the last six months. |
| **Reques L et al. (2024)** | Euro Surveillance | France | Mpox in children and adolescents and contact follow-up in school settings in | Retrospective study | 6 Cases were reported in 15-17 years adolescents, four reported to be heterosexual contact. Cases among children |
|  |  |  | greater Paris, France, May 2022 to July 2023. |  | and adolescents were common in French context. |
| **Hoffmann C et al. (2022)** | Deutsches Ärzteblatt International | Germany | Monkeypox in Germany. | Retrospective study | Out of the 301 cases, all were MSM, 46.7% co-infected with HIV and only 41% of cases didn't report history of STIs in the last six months. |
| **Abed Alah M et al. (2022)** | Journal of Infection and Public Health |  | The story behind the first few cases of monkeypox infection in non-endemic countries, 2022. | Case series | Cases reported in Portugal, Spain, the UK, France, Germany and Czech Republic reported behaviors such as MSM, close physical contact abroad and linked with sauna – gay friendly event, event in Grand Canary Islands and Darkland festival in Belgium. |
| **Salvo et al. (2023)** | Virology Journal | Italy | Clinical presentation of human monkeypox virus infection during the 2022 outbreak: descriptive case series from a large Italian Research Hospital. | Case series | All cases were reported to be MSM. Symptoms included vesicular rashes, genital rashes, lymphadenopathy and fever. More than half of the patients were co-infected with HIV-1. |
| **Gao S et al. (2024)** | International Journal of Infectious Diseases | Europe and America | Global transboundary transmission path and risk of Mpox revealed with Least Cost Path model. | Modelling study | The Least Cost Path (LCP) method identified facilitation of population migration by strongly established travel networks especially from London, England, to Spain and the States. |
| **Orviz et al. (2022)** | Journal of Infection | Spain | Monkeypox outbreak in Madrid (Spain): Clinical and virological aspects. | Case series | Though most of the patients were MSM, cases have been reported in homosexual/ bisexual IV drug users and heterosexuals. Risk factors include people living with HIV, unprotected oral sex, unprotected anal intercourse and unprotected vaginal sex. Only one-fourth were vaccinated for smallpox in childhood. |
| **Ianache I et al. (2024)** | Travel Medicine and Infectious Disease | 4 CEE countries, 8 from European Union (EU), and 6 outside the EU | Mpox across countries from Central and Eastern Europe - 2022 outbreak | Review paper (Survey) | Most of the patients were male, median age 35, through sexual contact and were MSM. Patients reported high risk behaviors namely unprotected sex and MSPs and living with HIV. Syphilis, gonorrhea, and chlamydia were the most frequent STIs, diagnosed both before and simultaneously with Mpox. |
| **Shang W et al. (2023)** | Journal of Medical Virology |  | Spatiotemporal cluster of mpox in men who have sex with men: A modelling study in 83 countries. | Modelling study | Between May 1, 2022, and March 31, 2023, 12 spatiotemporal clusters of Mpox cases were found in MSM population and the most likely cluster was in Spain. |
| **Pisano et al. (2024)** | Travel medicine and infectious diseases | Italy | The never-ending story of mpox epidemic: Tracing a new cluster in Florence, Italy | Case report | MSM cluster of 17 cases was identified in Florence but the interlinking of cases wasn't discussed. |
| **Grothe, J H et al. (2023)** | Enfermedades Infecciosas y Microbiología Clínica | Europe | Monkeypox in children and adult women in Europe: Results from a flash VACCELERATE pilot survey | Survey-based descriptive study | Out of 88 institutions, 5 reported confirmed MPXV cases. Spain and Belgium had the highest numbers of infected women, while Belgium and the UK had a higher number of children infected. |
| **Garcia, de Rio et al. (2022)** | The Lancet Infectious Diseases | Spain | Monkeypox outbreak in a piercing and tattoo establishment in Spain | Descriptive case series | Poor hygiene and aseptic conditions were reported in a tattoo parlor, with a high viral load found on tweezers. |
| **Martinez, V et al. (2023)** | New England Journal of Medicine | Spain | MPXV Transmission at a Tattoo Parlor | Case report | 36% of parlor customers became infected, mostly females. |
| **Gonzalez, G et al. (2024)** | Eurosurveillance | Ireland | Multiple introductions of monkeypox virus to Ireland during the international mpox | Modelling study | Importation of cases into Ireland from countries such as the UK, Portugal, Germany, Switzerland, and both North |
|  |  |  | outbreak, May 2022 to October 2023 |  | and South America. Ireland exported cases to Spain, Portugal, Slovakia, Italy, and Belgium. Genetic sequencing suggested that cases in Ireland were linked to those in Portugal. |
| **Wurtzer, S et al. (2022)** | medRxiv | France | First detection of Monkeypox virus genome in sewersheds in France | Surveillance study | MPXV was found in wastewater in France, affecting three out of sixteen sewersheds, indicating widespread community infection. |
| **Vaughan, A et al. (2018)** | Eurosurveillance | United Kingdom | Two cases of monkeypox imported to the United Kingdom, September 2018 | Case report | Two MPXV cases were reported in September 2018 in the UK. One involved a Nigerian naval officer, and another was linked to travel in Nigeria. |
| **Caria, J et al. (2022)** | Infectious disease reports | Portugal | Clinical and Epidemiological Features of Hospitalized and Ambulatory Patients with Human Monkeypox Infection: A Retrospective Observational Study in Portugal | Retrospective observational study | 59.5% of cases involved people living with HIV, with most of them being MSM. HIV patients tended to be older and had recently traveled internationally. |
| **Vanhamel, J et al. (2022)** | Sexually Transmitted Infections | Belgium | Understanding sexual transmission dynamics and transmission contexts of monkeypox virus: a mixed-methods study of the early outbreak in Belgium (May–June 2022) | Mixed-methods study | 28% of cases were linked to attendance at gay-oriented festivals, with some reporting sexual activity at events such as the Fetish Festival in Belgium and Pride Festivals in Spain. |
| **Gazecka, M et al. (2023)** | International Journal of Infectious Diseases | Poland | Mpox virus detection in the wastewater and the number of hospitalized patients in the Poznan metropolitan area, Poland | surveillance study | In Poznan, cases were linked to wastewater, indicating spread among the population. |
| **Ramírez-** | Open Forum | Spain | Clinical and Epidemiological | Cohort study | Most cases involved MSM, though transmission between |
| **Olivencia, G et al. (2024)** | Infectious Diseases |  | Characteristics of the 2022 Mpox Outbreak in Spain (CEME-22 Study) |  | other groups was recorded (male-to-female, female-to-male, and female-to-female). Median sexual partners were three, with 39% living with HIV. A small percentage had received smallpox vaccinations, and one patient died. |
| **Cassir, N et al. (2022)** | Emerging Infectious Diseases | France | Observational Cohort Study of Evolving Epidemiologic, Clinical, and Virologic Features of Monkeypox in Southern France | Observational cohort study | 92% MSM out of the reported 136 cases and 4% heterosexual cases with HIV coinfections being a risk factor |
| **Aguilera-Alonso, D et al. (2022)** | The Lancet Child and Adolescent Health | Spain | Monkeypox virus infections in children in Spain during the first months of the 2022 outbreak | Case series | Cases included four children younger than 4 years and twelve adolescents aged 13-17 years. Cases in younger populations were primarily due to household and contaminated material exposure. |
| **Girometti, N et al. (2022)** | The Lancet Infectious Diseases | UK | Demographic and clinical characteristics of confirmed human monkeypox virus cases in individuals attending a sexual health center in London, UK: an observational analysis | Observational analysis | All 54 cases were MSM with median age of 41 years and 24% living with HIV. |
| **Martínez, J I et al. (2022)** | Eurosurveillance | Spain | Monkeypox outbreak predominantly affecting men who have sex with men, Madrid, Spain, 26 April to 16 June 2022 | Descriptive case series | 99% of the cases were Men, 44.3% with HIV co-infection, 11% of the cases were on PrEP. |
| **Krug, C et al. (2023)** | Eurosurveillance | France | Mpox outbreak in France: epidemiological characteristics and sexual behaviour of cases aged 15 years or older, 2022 | Descriptive study | 97% of the total 4812 cases were men, 3% were women. Transmission through sexual contact - 71.1%, household contact - 16.8%. 47.9% attended MSM venues, 24.8% living with HIV and 18.4% were smallpox vaccinated. 96% were MSM, 70.7% reported more than 2 sex partners, |
|  |  |  |  |  | 77.2% reported anal sex and 39.4% reported chemsex behaviors. |

## Results

The initial search yielded 570 studies. After removing 149 duplicates, 421 studies were screened. Of these, 326 studies were excluded based on title and abstract screening. The remaining 86 studies were assessed for full-text eligibility, with 9 studies excluded due to the lack of full text. Eventually, 38 studies were included in the final analysis.

### Study characteristics

The final analysis included 38 studies conducted across various European countries, with specific representation from Ireland (n=1), Spain (n=7), Portugal, Belgium (n=2), Poland, Germany (n=4), Sweden, the Netherlands, Italy (n=3), France (n=4), Malta, the UK (n=3), broader European Region (n=3), and one study across four Central and Eastern European (CEE) countries (eight studies were conducted within the European Union (EU), covering the Czech Republic, Romania, Poland, Greece, Hungary, Croatia, Bulgaria, and Slovakia, while six were conducted outside the EU, including Serbia, Bosnia-Herzegovina, Moldova, Turkey, and Multicountry studies within the EU). These studies employed diverse methodologies, including review studies, retrospective analyses, cohort studies, narrative reviews, modelling studies, case reports, and descriptive case series.

**Figure 2.**
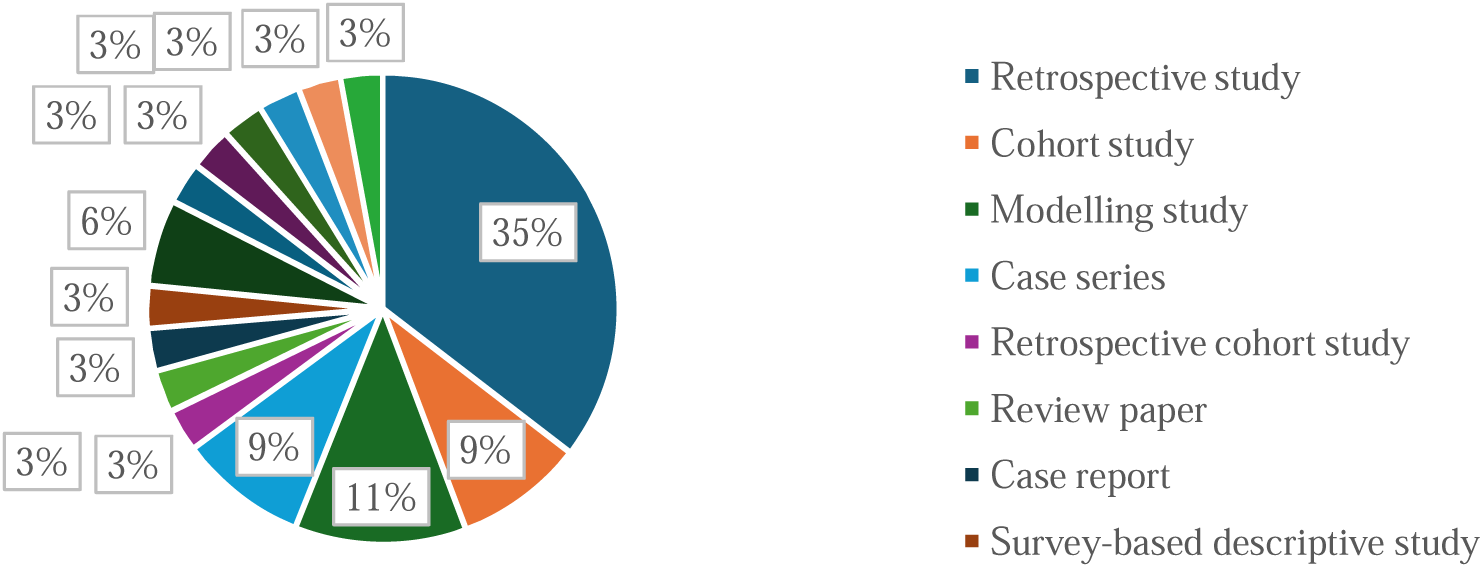
Study Designs included in the review.

**Figure 3.**
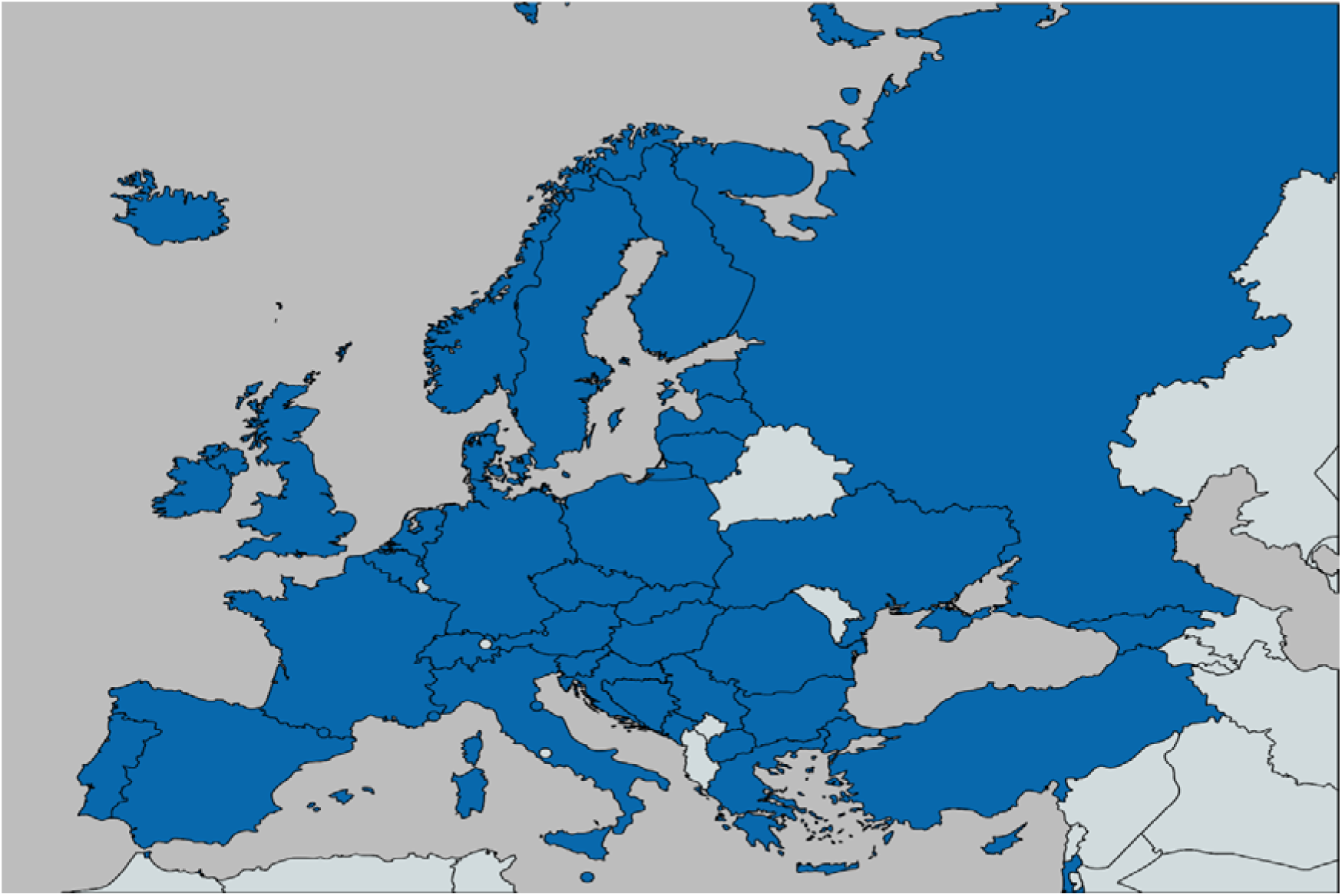
Study Settings across Europe in the review.

The review identified two key themes: Epidemiology and Risk Factors/Behaviors, along with various sub-themes under Risk Factors/Behaviors (see Figure 4).

**Figure 4.**
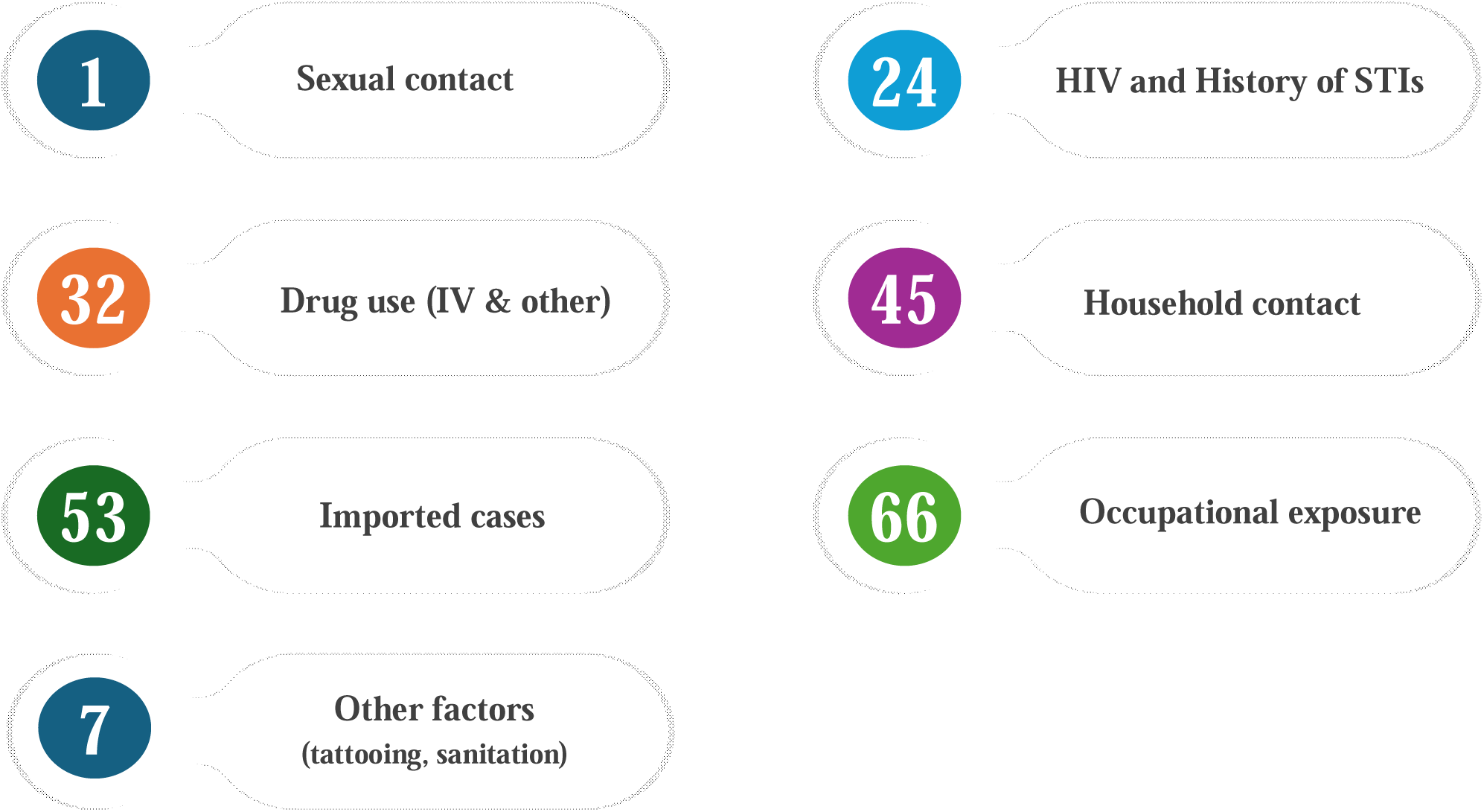
Risk factors/behaviors associated with Mpox in Europe.

### A. Epidemiology in Europe

The total number of confirmed and suspected human mpox cases worldwide from 1970 to July 1, 2022, was 46,915, with endemic regions accounting for 2,805 confirmed cases and 38,327 suspected cases. However, between May 7, 2022, and July 1, 2022, 5,783 mpox cases were reported in 52 non-endemic nations, including Europe, the USA, Australia, and the Middle East. The most affected countries during this period were the United Kingdom (1,235 cases), Germany (1,054), Spain (800), France (498), the United States (459), Portugal (402), the Netherlands (288), Canada (287), Italy (192), Belgium (117), Switzerland (91), Israel (42), Ireland (39), Austria (37), Sweden (28), Brazil (21), and Denmark (20) (19).

While the incidence of MPXV is more common in men, cases in women were also reported, with Spain accounting for one-third of the mpox cases among women in Europe in 2022. Moreover, complications were reported to be higher in women than in men (p=0.019) (20).

Until July 2022, Europe remained the epicenter of the geographically widespread outbreak, with a steady increase in cases and the number of affected countries (21). Following July 2022, the number of cases declined rapidly and remained low until June 2023. However, in 2023, cases began to rise again, increasing from approximately 6.6 weekly cases during the first five months to 30.3 weekly cases during the remainder of the year. Despite this resurgence, the total number of cases during Period 2 (July 2023 to May 2024) (n=1,432) was significantly lower than in Period 1 (March 2022 to June 2023) (n=25,866). Furthermore, the geographic spread of the virus decreased from 41 countries to 25 countries.

Nonetheless, a notable resurgence was observed in Spain, Portugal, Germany, the UK, and France, which together accounted for 76% of cases during Period 2. Additionally, a demographic shift emerged, with younger age groups being more affected in Period 2 compared to Period 1 (22).

Vaughan et al. (2024) reported that 97% of the cases between 2023 and 2024 involved the global sequence MPXV Clade IIb, which is less virulent. The remaining sequences were associated with Clade IIa and Clade I lineages. The usual incubation period for the virus ranges from 2-3 to 20 days, with only 5% of cases exhibiting an incubation period longer than 21 days (23). The mean time from the onset of symptoms to clinical assessment was reported to be 7 days (24). The median time between onset of symptoms and healing was reported to be 19.5 days (25).

### B. Risk factors/Behaviors

The review identified several key themes related to risk factors for mpox transmission in Europe, including sexual contact, HIV status and history of sexually transmitted infections (STIs), drug use, household contact, imported cases, occupational exposure, and other factors.

#### 1. Sexual contact

##### a. Male to Male Sexual Contact

Mpox transmission is notably prevalent among men who have sex with men (MSM), including individuals involved in transactional sex for money or drugs (24–32). Vallejo-Plaza A et al. (2022) observed significant differences in transmission rates between men and women during sexual contact, with 92.9% of male cases versus 65.7% of female cases (p<0.001) (20).

In Belgium, an epidemiological study revealed that 86% of mpox cases were identified as MSM, with sexual contact being the primary mode of transmission rather than other person-to-person interactions (33). Similarly, the first two mpox cases reported in Germany were linked to unprotected homosexual intercourse, where one of the patients was a MSM sex worker (34).

A significant outbreak was reported in Berlin during May-June 2022, where many cases attended events such as Club C in Berlin, the Gay Pride Festival in Grand Canaria, and a fetish festival in Antwerp (23). This might be linked to the case series in Italy which identified four mpox cases in May 2022 among young men, three of whom attended a mass event in the island of Grand Canary, while one was involved in providing sexual services with various male partners (35).

Interestingly, no widespread transmission was detected within Dutch MSM sexual networks prior to May 2022. Of 410 samples tested, 399 were negative, indicating a lack of undetected extensive transmission in the Netherlands before May 2022 (36). However, in England, 96% of cases reported on May 25, 2022, were linked to the Gay, bisexual, and other men who have sex with men (GBMSM) sexual networks. A survey conducted with those positive cases in England reported that 98% of the cases identifying as gay or bisexual. Additionally, 64% of these cases were associated with sex on-premises venues, festivals, cruising grounds, and private sex parties (37). Similar patterns of transmission were observed in Spain, Italy, France, Germany, the Netherlands, Belgium and Czech Republic (26, 38, 39).

From March 7 to August 23, 2022, across 41 countries in the WHO European Region, 93.9% of reported mpox cases were infected through sexual contact. Additionally, a significant proportion (98.8%) of the reported cases were male with a median age of 37. Among these, 96.9% of the male cases were MSM (21). A Multicountry study indicated that 98% of mpox cases reported in 2022 and 2023 were male, predominantly MSM (96%), aged 31-40 years, and transmitted through sexual contact (94%) (22). Shang et al. (2023) identified 12 spatial-temporal clusters of mpox cases in MSM populations across regions including Spain, Western Europe, and North America, further underscoring the significance of this transmission route (40). Similar clusters were also reported in Italy (41). A study encompassing four CEE countries, and eight European nations found that 99.3% of the cases were male, 90.2% were MSM, 65.5% reported unprotected sex, and 39.6% had multiple sexual partners. The study also highlighted significant differences in transmission patterns between European and non-European cases (MSM: p<0.0001; chemsex: p=0.008; group sex: p=0.005; multiple partners: p=0.025) (42). Similar trends were reported in a retrospective cohort study in Malta (43).

##### b. Heterosexual contact

While the majority of mpox cases occur among MSM, heterosexual transmission has also been documented (30, 44). Orviz et al. (2022) reported that 2.1% of cases in their study involved men who only had sexual contact with cisgender women, with 4.1% involving unprotected vaginal sex (24). In Buenos Aires, 3.8% of the cases reported in May 2022 were identified as heterosexuals (27). Additionally, Germany reported an mpox case in June 2022 involving a man who had sexual contact only with an unknown female sex worker (38). France documented six mpox cases among individuals aged 15-17 years between July and September 2022; four of these cases involved sexual contact, but none were identified as MSM, supporting the possibility of heterosexual transmission (45).

#### 2. HIV and History of STIs

There is a strong association between mpox cases and individuals with HIV or a history of STIs, with syphilis, gonorrhea, and chlamydia being the most reported infections (25, 30–32, 46, 47). Recent STI diagnoses within the last six months, particularly gonorrhea, were prevalent among mpox patients (38). Studies have shown that HIV prevalence among mpox cases is significantly higher than the general population, with notable differences between men and women (20). Additionally, the mean age of mpox patients with HIV tends to be higher compared to those without HIV (29).

The first mpox case reported in Germany involved a person living with HIV (34). In June 2022, 46.7% of mpox patients in Germany were HIV-positive, 44.7% were on pre-exposure prophylaxis (PrEP), and only 41% had not been diagnosed with any STIs in the past six months (28, 38). Similarly, a retrospective cohort study in Malta found that 27% of mpox cases involved individuals living with HIV (43). Across Europe, from March 7 to August 23, 2022, 37.2% (3,070/8,257) of reported mpox cases were among people living with HIV (21).

In Belgium, 23.2% of HIV-PrEP users were classified as “central network nodes” due to their Treponema pallidum antibody positivity (TPA-positivity) and higher number of sexual partners, although they were not more likely to have Orthopoxvirus-specific antibodies compared to TPA-negative HIV-PrEP users (33). Among the 10 mpox-related deaths reported from 2023 to 2024, eight were HIV-positive (22). Ianeche et al. (2024) showed history of STIs (p=0.004) and HIV infection (48.7%, p<0.0001) were common among mpox cases, with significant differences between European and non-European cases (HIV: p=0.042) (42). An Italian study reported that 56.25% of patients with mpox were HIV-1 coinfected (25). Orviz et al. (2022) found that 39.5% of their study’s mpox cases were living with HIV, and 47.9% of those were on daily PrEP with TDF/FTC (24). Similar findings were reported by Candela et al. (2023) in Italy and Hoffman et al. (2023) in Germany, with HIV prevalence in 47% and 46.9% of mpox cases, respectively.

#### 3. Drug use (IV & other recreational substances)

Drug use, particularly intravenous (IV) and recreational drugs, has also been identified as a risk factor. In a study conducted during the Madrid outbreak, 2.1% of participants were MSM who were IV drug users (24). An epidemiological study at a tertiary hospital in Italy between May and October 2022 reported recreational substance use, including smoking (37%), alcohol (39%), and chemsex drugs such as methamphetamine, mephedrone, gamma-hydroxybutyrate (GHB), and gamma-butyrolactone (27%) among mpox patients (32, 46).

#### 4. Household contact

Between May 2022 and July 2023, 19 mpox cases were reported in children under 18 years of age in France. The transmission risk among these children was primarily linked to intrafamilial contact (45). Similar findings were reported by Grothe et al. (2023) highlighting the confirmed cases in adult women and younger children with highest cases reported in Belgium, UK and Spain (48). Additionally, cases in children in Spain were attributed to household contact (49).

#### 5. Imported cases

Imported cases of mpox pose a significant risk for spreading the disease, particularly among displaced populations.

Kinoshita et al. (2023) highlighted the risk of global importation of mpox cases, especially in locations with large populations and well-connected flight networks, such as London, UK (50). This risk is underscored by two imported cases reported in the UK in September 2018, where both patients had traveled from Nigeria (51). Phylogenetic analysis in the Netherlands confirmed the introduction of several Clade II B.1 lineage clusters, (which is a global sequence) into the country, which then spread locally within the community (52). Similar patterns of importation and spread have been reported from Ireland to other countries and vice versa (53). Additionally, the Least Cost Path Model study supports the notion that population migration has been facilitated through strong travel networks, especially from London to Spain and the United States, facilitated the importation and transmission of mpox cases (54).

#### 6. Occupational exposure

Certain occupations present a higher risk of mpox infection. Healthcare professionals who come into contact with infected patients, veterinarians handling contaminated animals, dentists exposed to oral lesions and droplets, and zoo keepers or pet shop employees dealing with exotic or imported animals from endemic areas are particularly vulnerable (30).

#### 7. Other factors (e.g. animal contact, tattooing, poor sanitation practices)

Additional transmission routes include animal contact and non-sexual, non-mother-to-child pathways. An epidemiological study in Spain linked a tattoo studio to an outbreak of mpox cases among women, accounting for 8.9% of the cases (20, 55, 56).

Other transmission routes include person-to-person, fomite, and non-healthcare-related transmission. (21). Additionally, poor sanitation practices, including contamination via sewage systems, have been implicated in the spread of mpox. Cases linked to sewage systems have been reported in France, Poland, and other European countries (57, 58). Furthermore, contaminated materials have been associated with mpox cases in children in Spain (49).

## Discussion

### Summary of key findings

Mpox was first reported in Europe in 2020, with the initial case in the UK linked to travel history from Nigeria (12). By 2022, the disease had spread widely across Europe (19), affecting non-endemic countries primarily through person-to-person transmission, particularly via sexual contact. The cases were less deadly, as the spread was mainly associated with global sequences of Clade IIb, which is less virulent (23). Although MSM constituted the majority of cases (21, 24–28, 32, 36, 44), heterosexual transmission was also documented (24, 27, 32, 38, 44, 45). Cases have also been reported among women (20, 59). Individuals with multiple sexual partners were identified as being at higher risk (42, 43), with several cases linked to attendance at LGBTQ+ international festivals, parades, and saunas (26, 37–39).

Several studies revealed that nearly half of the reported cases involved individuals living with HIV, and a significant number had a history of STIs within the past six months (20–22, 25, 32–34, 38, 42, 43, 46, 47). Cases among IV drug users were also documented (24, 46). Frequent travel was identified as a significant factor in the spread of the disease (50, 52, 54), along with other risk behaviors such as intrafamilial contact (45, 49), animal contact (21, 60), occupational exposure (21) and tattooing (20, 55, 56). Notably, only a small percentage—less than 15%—of the affected individuals had received the smallpox vaccine during childhood (21, 24, 44, 46). The cessation of routine smallpox vaccination has been cited as one of the reasons for the increase in mpox cases (61).

To curb the spread of mpox, public health measures such as enhanced surveillance and monitoring (61–63), reducing the number of sexual partners (33), and considering smallpox vaccination (22) have been emphasized.

### Comparisons with existing literature

The high-risk behaviors identified in this scoping review—such as MSM activity, MSPs, and unprotected sex among promiscuous individuals and sex workers—are consistent with existing literature. Most studies have reported cases primarily in MSM and GBMSM populations, likely due to the extensive sexual networks within these communities (64, 65). The EMIS 2017 survey of European men who have sex with men found that the number of female sex partners ranged from zero to ten or more, and the number of male sex partners ranged from none to fifty or more (65). However, condom use among MSM and GBMSM was more consistent compared to heterosexuals during vaginal sex (66). This might suggest a potential shift in mpox cases from MSM networks to heterosexual networks, similar to the HIV burden transition observed between the 1990s and early 2000s (67).

Notably, similar risk behaviors, including MSPs and unprotected sex, have been reported among heterosexual men and women as well (68). MPXV has been isolated from semen at the onset of infection, with prolonged viral emission observed (69, 70). The presence of higher viral loads in rectal secretions (71, 72) exacerbates the disease burden among MSM and GBMSM. The significant disease burden in the LGBTQ+ community may be attributed to a combination of biological, behavioral, and sociocultural factors (73). Notably, individuals living with HIV and those on PrEP were also affected with mpox. This may be due to compromised immunity, which is common among HIV patients, making them more susceptible to mpox (74). While PrEP and antiretroviral therapies are designed to bolster immune defenses, studies suggest that immune recovery is not entirely complete even after HIV treatments, leaving these individuals vulnerable to MPXV (75). Contact with contaminated animals is one of the possible transmission routes with anthropozoonotic spillover demonstrated by a reported case of mpox transmission from a human to a domesticated dog in France (60). Mpox cases linked to contaminated wastewater and sewer sheds reported in France, Poland and other European Countries, consistent with findings from a study in Spain, where samples from wastewater treatment plants were found to be contaminated with MPXV (76).

In case of displaced population, people crossing borders daily to return to Ukraine are at heightened risk of contracting and spreading mpox, especially asymptomatic cases. These populations, often forced to leave their homes due to ongoing conflicts, inadvertently contribute to the spread of the virus in non-endemic regions (77).

This scoping review also noted a minimal number of mpox cases between 2023 and 2024 during Period 2, along with an increased rate of smallpox vaccination. This period predominantly involved clade IIb.1, which is less virulent. The cessation of routine smallpox vaccination is considered one of the contributing factors to the increase in mpox cases. However, experimental studies have demonstrated that smallpox vaccines are effective against mpox (78–80), and it is estimated that achieving collective immunity of at least 50.25% is necessary to halt the circulation of MPXV (81).

Studies mentioned hostile environment created by Eastern Europe towards the LGBTQ+ community has likely hindered the implementation of widespread, appropriate public health measures and discouraged individuals from seeking clinical examination and treatment (77). This contrasts with the 1981 AIDS epidemic in the United States, where major public health initiatives, funding, and community activities were spearheaded by the LGBTQ+ community, particularly in cities like San Francisco and New York (82).

A 2023 modelling study investigating the reproduction number of mpox in Europe revealed that the average reproduction number in Spain, France, Germany, the UK, the Netherlands, Portugal, and Italy was 2.04 at the start of the outbreak in 2022. This number dropped below 1 by the end of the outbreak, indicating a significant reduction in mpox cases and suggesting a low likelihood of future outbreaks in Europe as transmission chains were disrupted (83). This aligns with the observation that most cases in Europe during 2022-2023 were caused by the Clade II lineage, which is less virulent. However, the PHEIC declared in August 2024 was due to an increase in cases in the DRC, where the more virulent Clade I lineage is prevalent, raising concerns about potential importation to Europe (3, 13, 14).

### Methodological strengths and limitations

This review was conducted using Arksey and O’Malley’s six-step methodological framework (16), which provided a structured approach to examining the epidemiology of mpox. However, several limitations should be considered when interpreting the findings.

Firstly, while a comprehensive approach was employed, reliance on specific search criteria may have resulted in the exclusion of some relevant publications that met the inclusion criteria. Additionally, this review did not include an assessment of study quality, as the primary focus was on capturing the breadth of work related to the topic, rather than restricting the review to studies meeting specific scientific rigor standards.

Moreover, the review was limited to articles published in English and other languages (with translated version available online) within the last 10 years, potentially excluding important research published in other languages or prior to this time frame. These excluded studies might have provided key insights into the epidemiology of mpox in different regions or historical contexts. While the review encompassed Multicountry studies within the European Union, it did not involve a detailed analysis of all individual countries within Europe. This may limit the applicability of the findings to specific national or cultural contexts. However, the review captured the context in countries that experienced notable surges in cases.

Furthermore, there weren’t many studies reporting on the epidemiology of mpox after week 22 of 2024 were identified, highlighting a gap in the current literature that future research should aim to address. Despite these limitations, this review makes a significant contribution by providing a detailed overview of mpox epidemiology in Europe, including an exploration of risk behaviors that have not been widely studied in other papers.

### Implications for public health education, research and practice

To address the evolving epidemiology of mpox, it is crucial to expand research beyond the currently predominant focus on homosexual networks (84). Epidemiological and case-control studies should be encouraged for other risk groups, including MSPs among heterosexual networks, IV drug users and their sexual partners, and individuals engaging in sexual activities in exchange for money or drugs. Given that mpox is not strictly a sexually transmitted disease and can be transmitted through person-to-person contact, studies focusing on intrafamilial transmission should also be designed.

Since mpox is not currently screened for during blood transfusions and transplantations (85, 86), it is important to investigate potential cases of transmission through blood and blood products (87), particularly in hemophilia patients and transfusion cases, even though the typical incubation period is less than 20 days (23–25). Monitoring displaced populations and refugees is essential, as they may import cases, and appropriate health services should be provided to them. Mandatory mpox testing in refugee camps serves the purpose.

Smallpox vaccination campaigns should be conducted, and mandatory vaccination or testing for mpox should be implemented for individuals traveling to non-endemic countries, particularly those coming from endemic regions. Quarantine measures should be enforced for travelers displaying symptoms and contact tracing should be rigorously practiced. Flight networks from endemic countries could be temporarily reduced, and curfews implemented, if necessary, to limit the spread of the virus.

General practitioners (GPs) should actively promote vaccination among high-risk groups, such as GBMSM, sex workers, and IV drug users, during routine consultations. Public awareness campaigns should emphasize safe sex practices, including the use of condoms, dental dams, and other protective measures. The availability of high-quality condoms through free vending machines should be expanded, and the importance of reducing the number of sexual partners and limiting attendance at large gatherings should be stressed. Regular STI testing should be encouraged, and PrEP should be recommended for individuals in high-risk categories.

Studies on risk behaviors within GBMSM networks need to be carefully designed, with interventions crafted to respect the emotional, social, and moral factors of these communities. Behavioral interventions should focus on promoting clean needle practices, safe sex, and reducing risky behaviors to help prevent the spread of mpox. Community initiatives similar to successful programs, such as the Miami Community Outreach Project for Injecting Drug Users and Other High Risk Groups, should be designed and encouraged to address these risk groups (88). To limit the risk of transmission, the import of animals from endemic regions should be banned. Children should be advised to avoid contact with potentially infected species, particularly African rodents. Occupational safety measures must be strictly followed, with doctors, dentists, veterinarians, and zookeepers using appropriate PPE including gloves, masks, and respirators.

Improvements in hygiene and sanitation practices are essential, with a particular emphasis on regular handwashing and sanitizing in both household and business environments, especially in settings like tattoo parlors and salons, where equipment must be properly sterilized. Wastewater treatment plants and water systems should be continuously monitored, sanitized, and placed under strict surveillance to prevent contamination. Patients and caregivers should frequently sanitize their hands and any potentially contaminated surfaces. Infected individuals should quarantine and seek appropriate medical care from general practitioners and specialists to limit further transmission.

## Data Availability

All data produced in the present work are contained in the manuscript

## Acknowledgments

I would like to acknowledge that there are no additional contributors or external support for this work.

## Notes

### Competing Interest Statement

The authors have declared no competing interest.

### Funding Statement

This study did not receive any funding

### Summary of Updates

Added another author, amended a few redundancies

